# App-Guided Hepatitis Self-Testing Versus Provider-Administered Rapid Testing Among Out-of-School Young Adults in Ibadan, Nigeria

**DOI:** 10.64898/2026.09.12.26362907

**Authors:** Adekunle O. Adeluwoye, Yetunde M. Rufai, Adebusayo R. Olayinka, Naomi N. Adeluwoye, Ayanfeoluwa Alabetutu

## Abstract

**Background:** Hepatitis B virus (HBV) seroprevalence reaches 10–12% among out-of-school youths in urban Southwest Nigeria, yet fewer than three in ten in this demographic have ever undergone HBV testing. E-health self-testing applications offer an accessible alternative delivery modality for hepatitis screening, but direct evidence comparing their acceptance against traditional laboratory-based delivery in informal commercial settings is lacking.

**Objective:** To determine whether e-health app–guided hepatitis self-testing achieves equivalent disease detection and superior acceptance outcomes compared with traditional laboratory-based screening among out-of-school youths in a high-footfall commercial site in Ibadan, Nigeria.

**Methods:** A cross-sectional parallel-arm pilot study recruited 120 out-of-school youths at commercial premises on Iwo Road, Ibadan (test group: n=60, e-health app-guided hepatitis B and C self-testing; control group: n=60, traditional laboratory-based testing using identical rapid diagnostic kits). Pre- and post-test structured questionnaires assessed technology readiness, behavioural intention, perceived ease of use, and adoption barriers. The Technology Acceptance Model (TAM) and Unified Theory of Acceptance and Use of Technology (UTAUT) provided the analytical framework. Group comparisons used chi-square and Fisher’s exact tests (α=0.05).

**Results:** HBV seroprevalence was 10.0% in the test group and 11.7% in the control group (χ²=0.087, p=0.769), confirming statistically equivalent detection across modalities. Hepatitis C virus was undetected in both groups. The test group demonstrated greater willingness to use e-health testing over traditional laboratory services (81.6% vs 70.0%), higher comfort with mobile health technology (75.0% vs 71.7%), and equivalent recommendation intent (93.3% vs 95.0%). Prior e-health service use was significantly higher in the test group (28.3% vs 13.3%; χ²=4.09, p=0.043). Anticipated ease of use (75.0% “very comfortable”) substantially exceeded experienced ease post-use (41.7% “very or extremely easy”). Privacy concern was the leading adoption barrier in both groups (30.0% vs 36.7%), outranking cost (25.0% vs 15.0%).

**Conclusions:** E-health self-testing achieves non-inferior hepatitis B detection compared with traditional laboratory delivery while generating comparable acceptance outcomes. The gap between anticipated and experienced ease of use identifies usability as the primary design target. Privacy-centred tool design, not cost reduction alone, is the critical condition for scaling hepatitis e-health self-testing among out-of-school youths in urban Nigeria.

## 1. Introduction

Viral hepatitis remains a major cause of preventable liver disease, and the largest burden of hepatitis B virus (HBV) infection is concentrated in regions where diagnosis and linkage to care remain incomplete. The 2024 global hepatitis report estimates that testing and treatment coverage remain substantially below elimination targets, particularly in low- and middle-income countries [1]. Nigeria is consistently classified as a high-burden setting, although published estimates vary by population, region, assay and period; meta-analyses nevertheless show persistent endemic HBV infection and substantial heterogeneity across Nigerian studies [2,3]. Knowledge of HBV, prior testing, and access to preventive services are also uneven in community populations, including traders and other informal-sector groups [4,5].

Young adults who are no longer in formal education can be difficult to reach through school-based health programmes. Work from markets and motor parks in southwest Nigeria has long documented unmet sexual and reproductive health needs among out-of-school young people, and more recent Nigerian studies describe barriers to care among street-involved and informal-sector populations [6–9]. These populations are not homogeneous risk groups, and being out of school should not itself be treated as an infection-risk marker. The public-health relevance is instead one of service access: individuals whose daily lives are organised around informal commercial work may have fewer routine points of contact with preventive testing services.

Mobile delivery offers one possible route around that access problem, but the Nigerian digital environment should not be simplified to universal smartphone readiness. Recent GSMA analyses emphasise a persistent usage gap even where mobile broadband coverage is available, with affordability, device ownership, skills, safety and relevant content all influencing meaningful connectivity [10]. Nigeria is simultaneously expanding national digital-health governance and infrastructure [11], while WHO guidance cautions that digital interventions should strengthen rather than substitute for functioning health systems [12]. Systematic reviews similarly show that mHealth can improve selected health behaviours and service processes, but effects depend on design, context and implementation rather than mobile delivery alone [13–15].

Self-testing provides a particularly relevant comparator because it moves some steps of testing from a professional setting to the individual. Evidence is strongest for HIV self-testing, but multi-infection self-testing studies have also evaluated combined rapid testing for HIV, HBV and HCV [16,17]. HIV self-testing studies demonstrate high acceptability in many settings and show that distribution models can increase testing uptake, while also making linkage to confirmatory care a central implementation requirement [18–20]. Nigerian youth studies add locally relevant evidence: cost, testing method, privacy, preferred access location, support after a reactive result and youth-friendly delivery all influence uptake [21–25]. These findings support digital or self-testing approaches as plausible service-delivery options, but they do not establish that self-administered testing has the same diagnostic performance as professional testing for an individual patient.

The Technology Acceptance Model (TAM) and Unified Theory of Acceptance and Use of Technology (UTAUT) offer useful language for understanding why a technically available tool may or may not be adopted. TAM centres perceived usefulness and perceived ease of use as determinants of intention [26], whereas UTAUT adds performance expectancy, effort expectancy, social influence and facilitating conditions [27]. Health-informatics reviews show extensive use of these models, but also considerable variation in how constructs are operationalised and validated [28–30]. Privacy concerns can influence adoption alongside effort expectancy and self-efficacy [31]. In the present study, TAM and UTAUT were not administered as prospectively validated psychometric scales; they are therefore used as interpretive frameworks rather than as latent-variable models.

This pilot study compared an app-guided hepatitis rapid-testing pathway with provider-administered rapid testing among self-identified out-of-school young adults working around Iwo Road, Ibadan. The study describes observed HBsAg and HCV antibody results, digital readiness, willingness, experiential usability and self-reported barriers, and estimates the precision of selected between-arm differences. Because allocation was non-random, the arms were imbalanced at baseline, and no equivalence margin or within-participant reference standard was specified, the study does not test diagnostic equivalence or noninferiority. Usability, acceptance, and implementation feasibility of the app-guided pathway are the primary focus; diagnostic performance and the comparative effectiveness of the two delivery modalities are addressed separately as directions for future evaluation.

## 2. Methods

### 2.1 Study design, setting, and reporting approach

We conducted a cross-sectional descriptive parallel-arm pilot study between September and November 2023 at commercial premises on Iwo Road, Ibadan, Oyo State, Nigeria (approximately 7.38°N, 3.90°E). Iwo Road is a high-traffic commercial area containing markets, retail shops and service businesses. Provider-administered testing for the control arm was conducted at the Ibadan North East Local Government Area Council Health Centre Laboratory, Iwo Road, Ibadan, Oyo State – a local government-run primary health facility within the same commercial catchment area – while app-guided testing in the intervention arm was conducted at the point of recruitment (Figure 1). This report follows STROBE principles for transparent reporting of observational data [32] and draws on guidance for non-randomised pilot and feasibility studies [33–35]. The design is a non-randomised pilot, and feasibility-oriented reporting conventions are applied accordingly.

**Figure 1.**
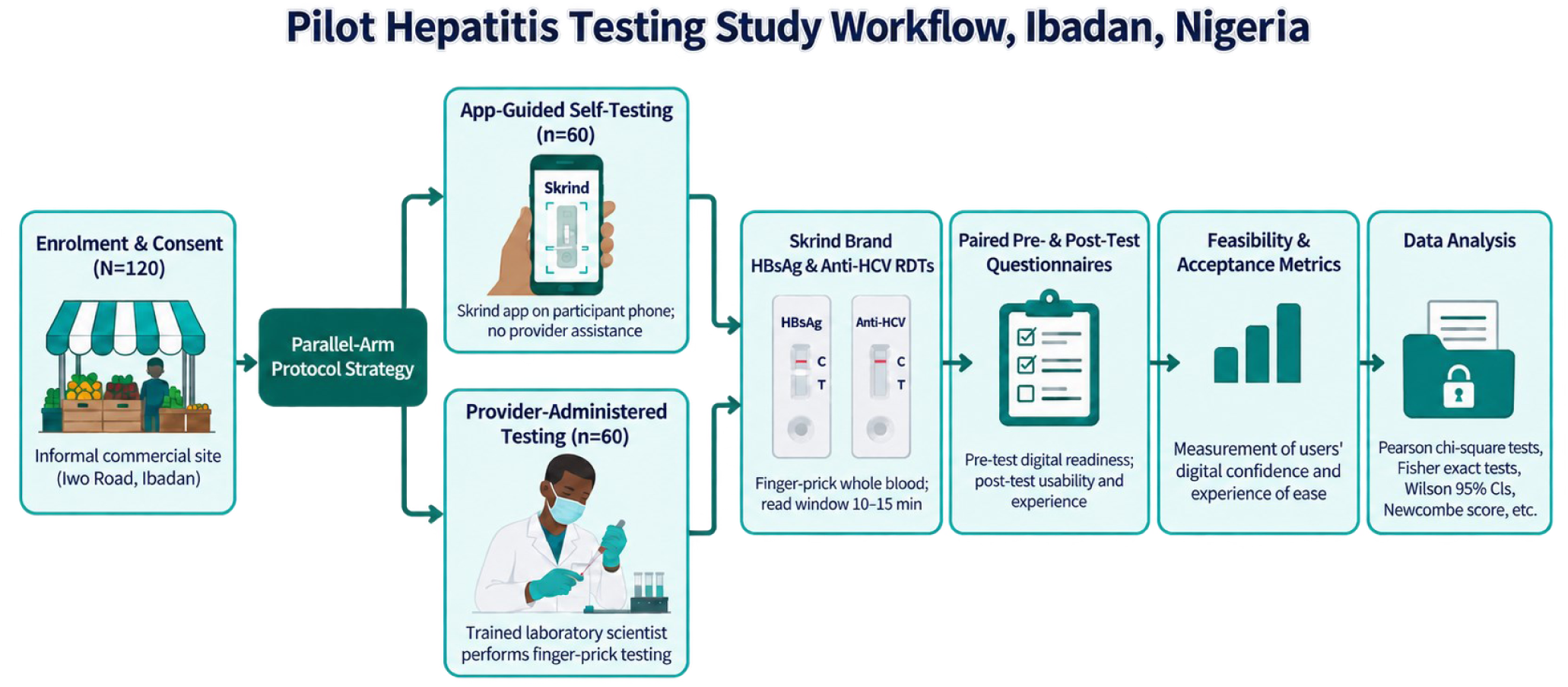
Study design and participant pathway. Participants were enrolled and allocated to app-guided self-testing (n=60) or provider-administered testing (n=60) using the same rapid diagnostic test brand for hepatitis B surface antigen and anti-hepatitis C virus antibody. Both arms completed paired pre- and post-test questionnaires assessing feasibility and acceptance, followed by descriptive statistical analysis.

### 2.2 Participants, recruitment, and allocation

Eligible participants were adults aged 18 years or older who self-identified as out of school and were present at the commercial recruitment site during the study period. The phrase out of school referred to current educational participation, not to lifetime educational attainment; several participants had completed tertiary education. Recruitment used convenience sampling. Participants entered the app-guided or provider-administered arm according to temporal recruitment blocks rather than a random allocation sequence. Pre-enrolment recruitment metrics (the number of individuals approached, screened for eligibility, or declining participation) were not systematically logged during data collection and cannot be reported. All 120 participants who provided informed consent completed both the rapid testing procedure and the paired pre- and post-test questionnaires; the analysed sample is therefore identical to the enrolled and consented sample, with no post-consent loss to follow-up to report.

The sample size was set at 60 participants per arm (N=120) for logistical and financial reasons because each app-guided participant required a rapid diagnostic test kit. No formal a priori power calculation was conducted. Pilot-study sample-size literature supports linking preliminary sample sizes to feasibility aims and precision rather than treating conventional hypothesis-testing power as the principal justification [36]. Post-hoc power calculations were not used, as they add little information beyond the effect estimate, sample size, and confidence interval already reported [37]. Ninety-five percent confidence intervals are reported throughout to convey the precision of the pilot estimates.

### 2.3 Testing pathways

Both arms underwent rapid testing for hepatitis B surface antigen (HBsAg) and antibody to hepatitis C virus (anti-HCV) using the same brand of commercially available immunochromatographic rapid diagnostic tests. In the app-guided arm, participants used an e-health mobile application (Skrind, version 1.0.4) on their own smartphones to follow test instructions, interpret the rapid-test result and record the outcome. In the provider-administered arm, a trained laboratory scientist performed the finger-prick rapid testing. Provider-arm participants were referred to and physically attended the Ibadan North East Local Government Area Council Health Centre Laboratory, Iwo Road, Ibadan, Oyo State, where testing was conducted by facility-based laboratory personnel. The contrast was therefore delivery modality, not assay chemistry. App-guided participants performed specimen collection, testing, and result interpretation entirely independently using the mobile application; no staff assistance was provided with lancet use or any other step of the app-guided procedure.

The rapid diagnostic test kits (distributed under the Skrind brand) were manufactured by Gem Medic Technology Co., Ltd, an affiliate of the GEMC Group. Both arms used whole blood specimens; app-guided participants self-collected a finger-prick specimen following in-app and leaflet instructions: selecting the index or middle finger, gently massaging the finger, cleaning it with an alcohol swab, pricking with the provided retractable lancet, applying the blood droplet to the sample space, and adding two drops of buffer fluid as indicated on the kit. The e-health application (Skrind, version 1.0.4) was used with the Skrind self-test kits for guided test administration and digital result interpretation. Participants were instructed within the app to start the timer in the Skrind app immediately after adding single-use buffer and to wait before reading the result; the primary instruction specified 15 minutes. Results were read by activating the smartphone camera and capturing the circled test region for app-based interpretation.

WHO testing guidance treats a reactive screening result as the beginning of a diagnostic and linkage pathway rather than as a complete endpoint [38]. Rapid HBsAg tests can have high specificity but sensitivity varies by product and clinical context [39,40]. Independent performance-validation data and published field sensitivity/specificity were available for these kits from the manufacturer. For the HBsAg rapid test kits used in this study, reported performance was high (sensitivity: 99.97%, specificity: 99.84%, accuracy: 99.90%), and for the HCV antibody rapid test kits, performance was similarly strong (sensitivity: 99.49%, specificity: 99.22%, accuracy: 99.31%). However, WHO prequalification status was not. Results are therefore reported as RDT-reactive rather than as confirmed chronic HBV or HCV infection.

While the Skrind mobile app (version 1.0.4) has care linkage to teleconsultation and advance laboratory testing that participants can utilize, all participants with a reactive HBsAg or anti-HCV result were counselled on the result and linked to a healthcare provider for confirmatory laboratory testing and further clinical evaluation for this study, providing a documented pathway from a reactive point-of-care result to advanced diagnostic and clinical care.

### 2.4 Questionnaires and TAM/UTAUT mapping

Structured paper questionnaires were administered before and after testing. The pre-test instrument captured sociodemographic characteristics, smartphone ownership and app-use patterns, prior e-health service use, disease knowledge, prior screening and perceived barriers. The post-test instrument was administered after the testing encounter. App-guided participants answered from direct experience; provider-arm participants were shown a demonstration of the e-health tool and therefore answered hypothetical questions about e-health use. The primary usability analysis is restricted to experiential app-arm responses.

Questionnaire items were mapped to TAM and UTAUT constructs to provide an interpretive framework for the acceptance and usability findings, consistent with common practice in early-stage mHealth pilot research [28–30]. As the constructs were not measured using a validated multi-item psychometric scale, the terms perceived usefulness, ease of use, behavioural intention, and facilitating conditions are used descriptively rather than as a basis for construct-level causal inference. A small number of post-test items referencing infectious diseases outside the study scope were excluded from the hepatitis-focused analysis.

### 2.5 Outcomes

Testing outcomes were the proportions with HBsAg-reactive and anti-HCV-reactive RDT results in each arm. Acceptance outcomes included stated willingness to use e-health testing, recommendation intention, baseline comfort with mobile health technology, and the perceived importance of access to testing. Experiential usability was defined using the app-arm post-use item rating the tool as very easy or extremely easy. Adoption-barrier items allowed multiple selections; privacy, convenience, cost and trust in tool accuracy were summarised separately. Because the pre-use comfort item and post-use ease item use different wording and response constructs, the difference between their percentages is reported as a descriptive contrast rather than as a paired change score.

### 2.6 Statistical analysis

The analysis was conducted in IBM SPSS Statistics version 20. Analyses use the reported counts and two-sided Pearson chi-square tests for selected 2×2 comparisons; Fisher exact tests was preferred where expected counts are small. For pilot interpretation, point estimates and precision are emphasised over significance testing. Wilson 95% CIs are reported for proportions, and Newcombe score intervals without continuity correction are reported for absolute differences between independent proportions [41]. No multiplicity adjustment was prespecified, so P values for secondary acceptance outcomes are exploratory. Because no participant-level dataset was available to fit an age-, smartphone-ownership-, or prior-use-adjusted model directly, the principal between-arm acceptance comparison was additionally examined using E-value sensitivity analysis [62]. The E-value estimates the minimum strength of association, on the risk-ratio scale, that an unadjusted covariate such as age group, smartphone ownership, or prior e-health use would need to have with both arm assignment and the outcome to fully explain an observed association, given the covariates already measured. This method uses only the reported point estimate and confidence interval.

No equivalence or noninferiority margin was prespecified, and each participant was tested through only one delivery pathway rather than through both modalities against an independent reference standard. A non-significant between-arm comparison therefore cannot demonstrate diagnostic equivalence or noninferiority [42]. Because the marked baseline age imbalance could not be addressed through stratification, regression, or propensity methods at the individual level, comparative acceptance estimates are interpreted as descriptive and potentially confounded. An E-value sensitivity analysis was performed to quantify how strong such confounding would need to be to account for the observed acceptance difference.

## 3. Results

### 3.1 Participant characteristics and baseline comparability

All 120 enrolled participants contributed testing outcomes and questionnaire data to the analysis. The two arms were not comparable on several baseline characteristics. Participants aged 18-35 years accounted for 88.3% (53/60) of the app-guided arm but 25.0% (15/60) of the provider arm, an absolute difference of 63.3 percentage points (95% CI 47.2 to 74.3; P<.001). Smartphone ownership was also higher in the app-guided arm (100%, 60/60) than in the provider arm (83.3%, 50/60), as was prior e-health use (28.3%, 17/60 vs 13.3%, 8/60). These imbalances are important because age and prior digital exposure are plausible determinants of both technology acceptance and ease of use. The full five-category age distribution is also reported in Table 1 for transparency.

**Table 1.** Baseline sociodemographic and digital-use characteristics by study arm.

| Characteristic | App-guided (n=60) | Provider-administered (n=60) | Difference, percentage points (95% CI) | P value |
| --- | --- | --- | --- | --- |
| Age 18-25 years | 26/60 (43.3%; 31.6%-55.9%) | 8/60 (13.3%; 6.9%-24.2%) |  |  |
| Age 26-35 years | 27/60 (45.0%; 33.1%-57.5%) | 7/60 (11.7%; 5.8%-22.2%) |  |  |
| Age 36-45 years | 5/60 (8.3%; 3.6%-18.1%) | 17/60 (28.3%; 18.5%-40.8%) |  |  |
| Age 46-55 years | 2/60 (3.3%; 0.9%-11.4%) | 14/60 (23.3%; 14.4%-35.4%) |  |  |
| Age 56+ years | 0/60 (0.0%; 0.0%-6.0%) | 14/60 (23.3%; 14.4%-35.4%) |  |  |
| Age 18-35 years (combined, summary) | 53/60 (88.3%; 77.8%-94.2%) | 15/60 (25.0%; 15.8%-37.2%) | +63.3 (+47.2 to +74.3) | <.001 |
| Female gender | 36/60 (60.0%; 47.4%-71.4%) | 31/60 (51.7%; 39.3%-63.8%) | +8.3 (-9.2 to +25.2) | .358 |
| Bachelor's degree | 34/60 (56.7%; 44.1%-68.4%) | 26/60 (43.3%; 31.6%-55.9%) | +13.3 (-4.4 to +30.0) | .144 |
| Smartphone ownership | 60/60 (100%; 94.0%-100%) | 50/60 (83.3%; 72.0%-90.7%) | +16.7 (+7.2 to +28.0) | .001* |
| Daily app use, multiple times/day | 45/60 (75.0%; 62.8%-84.2%) | 40/60 (66.7%; 54.1%-77.3%) | +8.3 (-7.9 to +24.0) | .315 |
| Prior e-health service use | 17/60 (28.3%; 18.5%-40.8%) | 8/60 (13.3%; 6.9%-24.2%) | +15.0 (+0.4 to +29.0) | .043 |
| Familiar with e-health HBV/HCV tools | 16/60 (26.7%; 17.1%-39.0%) | 13/60 (21.7%; 13.1%-33.6%) | +5.0 (-10.3 to +20.0) | .522 |

**Note:** Percentages were recalculated from integer counts. CIs for individual proportions are Wilson intervals; between-arm CIs are Newcombe intervals. P values are Pearson chi-square unless marked \* (Fisher exact). These comparisons are descriptive because allocation was not randomised.

### 3.2 Viral-hepatitis rapid-test outcomes

HBsAg was reactive in 10.0% (6/60; 95% CI 4.7%-20.1%) of app-guided participants and 11.7% (7/60; 95% CI 5.8%-22.2%) of provider-tested participants. The absolute difference was −1.7 percentage points (95% CI −13.5 to 10.1), and no statistically detectable between-arm difference was observed (Pearson χ²=0.086, P=.769). The confidence interval remains compatible with clinically meaningful differences in either direction, and the study design cannot estimate sensitivity, specificity or individual-level concordance between delivery modalities.

No participant had an anti-HCV-reactive result (0/120). The exact 95% CI for the overall observed proportion was 0%-3.0%, indicating that a low underlying prevalence remains compatible with the data.

**Table 2.** Rapid-test outcomes by delivery arm.

| Outcome | App<br>n/N | App % (95%<br>CI) | Provider<br>n/N | Provider %<br>(95% CI) | Difference, pp<br>(95% CI) | P value |
| --- | --- | --- | --- | --- | --- | --- |
| HBsAg<br>reactive | 6/60 | 10.0% (4.7%-<br>20.1%) | 7/60 | 11.7% (5.8%-<br>22.2%) | -1.7 (-13.5 to<br>+10.1) | .769 |
| Anti-HCV<br>reactive | 0/60 | 0% (0%-6.0%) | 0/60 | 0% (0%-6.0%) | 0.0 | Not<br>estimable |

*Note: HBsAg and anti-HCV results are described as RDT-reactive because confirmatory laboratory testing was not performed within this pilot. Overall anti-HCV result: 0/120, exact Clopper-Pearson 95% CI 0%-3.0%. Non-significance is not evidence of equivalence.*

### 3.3 Acceptance and experiential usability

Stated willingness to use e-health testing was high in both groups: 81.7% (49/60; 95% CI 70.1%-89.4%) in the app-guided arm and 70.0% (42/60; 95% CI 57.5%-80.1%) in the provider arm. The difference was 11.7 percentage points (95% CI −3.7 to 26.4; P=.136). Recommendation intention was similarly high (93.3%, 56/60 vs 95.0%, 57/60). Given the baseline age and digital-access imbalances, these between-arm contrasts should not be interpreted as an effect of delivery modality. To quantify how much unmeasured or unadjusted confounding by baseline differences such as age, smartphone ownership, or prior e-health use would need to explain the observed willingness difference, an E-value was calculated from the point estimate (risk ratio 1.17, calculated as 81.7%/70.0%). The E-value was 1.61, indicating that a covariate associated with both arm assignment and willingness by a risk ratio of at least 1.61-fold, beyond the variables already reported in Table 1, could account for the entire observed association; weaker associations could not. Given the magnitude of the observed baseline age imbalance (Table 1), an association of this size is plausible, and the E-value for the confidence interval is 1.0 because the interval already includes the null value. This formal sensitivity analysis is consistent with, and quantitatively reinforces, the descriptive interpretation already applied to between-arm acceptance comparisons in this pilot.

The more informative usability observation came from the app-guided arm alone. Before testing, 75.0% (45/60) described themselves as very comfortable with mobile health technology. After actual use, 41.7% (25/60; 95% CI 30.1%-54.3%) rated the tool as very easy or extremely easy. The 33.3-percentage-point difference is descriptive because the two questions measure related but non-identical concepts. It nevertheless shows that general digital confidence did not translate into uniformly high ease ratings for this specific testing workflow.

**Table 3.** Acceptance outcomes and app-arm experiential usability.

| Outcome | App-guided | Provider-administered | Difference, pp (95% CI) | P value | Interpretation |
| --- | --- | --- | --- | --- | --- |
| Very willing to use e-health testing | 49/60 (81.7%; 70.1%-89.4%) | 42/60 (70.0%; 57.5%-80.1%) | +11.7 (-3.7 to +26.4) | .136 | Between-arm; exploratory |
| E-value for willingness comparison (sensitivity analysis) | RR = 1.17; E-value = 1.61 | E-value for CI = 1.00 (CI includes null) | Not applicable | Not applicable | Quantifies confounding strength needed to explain the willingness difference; see Section 3.3 |
| Very comfortable with mobile-health technology (pre-use) | 45/60 (75.0%; 62.8%-84.2%) | 43/60 (71.7%; 59.2%-81.5%) | +3.3 (-12.4 to +18.8) | .680 | Baseline digital confidence |
| Willing to recommend e-health tool | 56/60 (93.3%; 84.1%-97.4%) | 57/60 (95.0%; 86.3%-98.3%) | -1.7 (-11.5 to +7.9) | .697 | Between-arm; exploratory |
| Testing access rated very important | 52/60 (86.7%; 75.8%-93.1%) | 44/60 (73.3%; 61.0%-82.9%) | +13.3 (-1.1 to +27.2) | .068 | Between-arm; exploratory |
| Tool rated very/extremely easy after actual use | 25/60 (41.7%; 30.1%-54.3%) | Not applicable in primary experiential analysis | Not applicable | Not tested | App-guided arm only |

*Note: Provider-arm hypothetical post-demo usability responses were excluded from the primary experiential usability analysis. P values are unadjusted and exploratory.*

### 3.4 Adoption barriers

Privacy was the most frequently reported barrier among the categories assessed: 30.0% (18/60) in the app-guided arm and 36.7% (22/60) in the provider arm selected privacy. Cost was selected by 25.0% (15/60) and 15.0% (9/60), respectively. Convenience was selected by 26.7% (16/60) and 25.0% (15/60). Few participants selected trust in tool accuracy (3.3%, 2/60 and 6.7%, 4/60), but this item should not be interpreted as a general measure of trust in digital health because it refers specifically to perceived diagnostic accuracy.

Figure 2 summarises the acceptance, usability, and barrier findings within the TAM/UTAUT interpretive framework.

**Figure 2.**
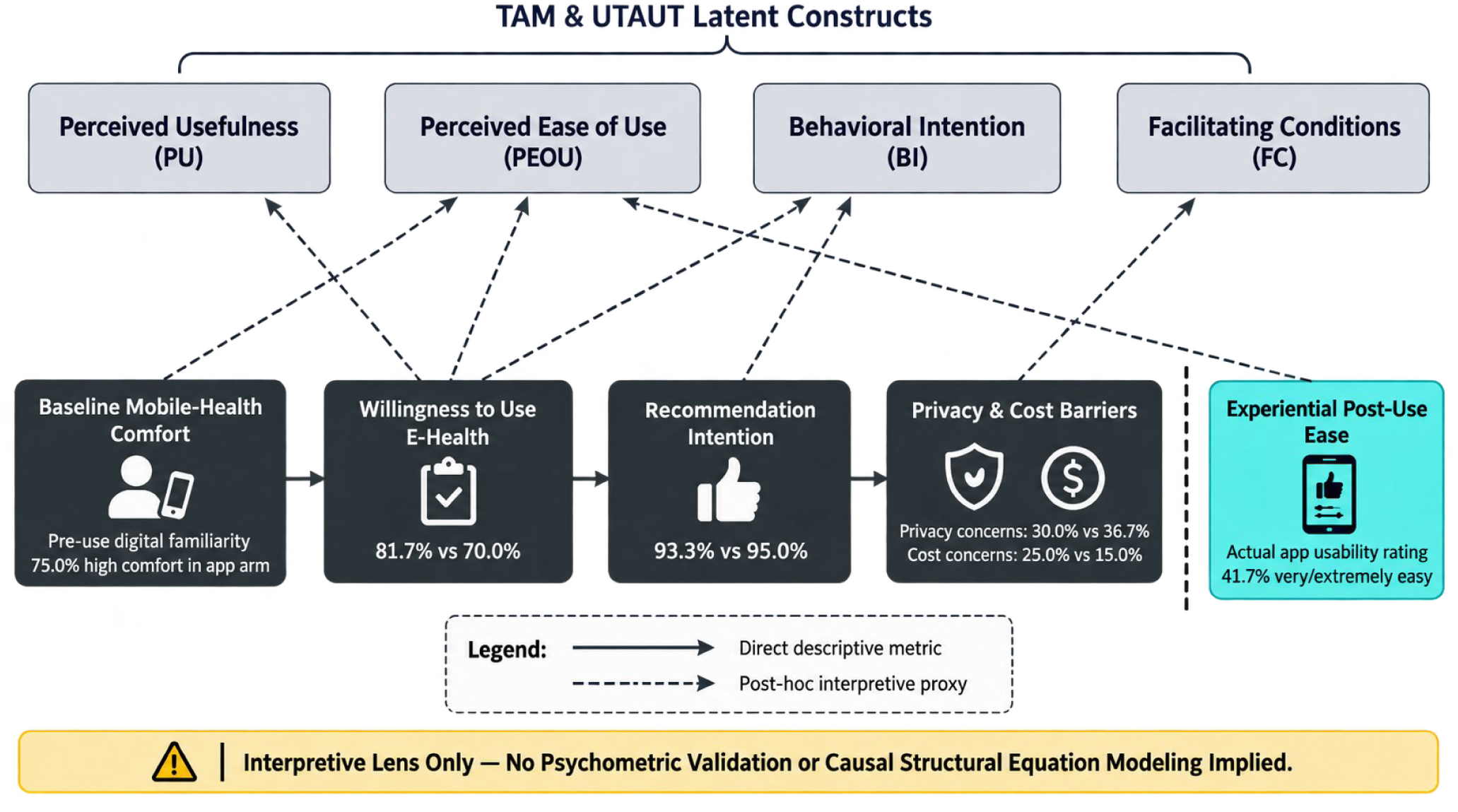
TAM/UTAUT-informed summary of acceptance, usability, and barrier outcomes. Directly measured descriptive items (solid arrows) and their post-hoc interpretive mapping to TAM/UTAUT constructs (dashed arrows) are shown alongside key observed values, including willingness to use e-health testing, recommendation intention, privacy and cost barriers, and experiential post-use ease. The constructs are applied as an interpretive lens only, without psychometric validation or causal structural modelling.

**Table 4.** Self-reported barriers to e-health hepatitis testing.

| Barrier category | App-guided (n=60) | Provider-administered (n=60) |
| --- | --- | --- |
| Privacy | 18/60 (30.0%) | 22/60 (36.7%) |
| Convenience | 16/60 (26.7%) | 15/60 (25.0%) |
| Cost | 15/60 (25.0%) | 9/60 (15.0%) |
| Trust in tool accuracy | 2/60 (3.3%) | 4/60 (6.7%) |

*Note: Participants could select more than one barrier, so percentages do not sum to 100%. No formal ranking or multiplicity-adjusted between-barrier test was prespecified. “Trust in tool accuracy” is kept separate from general technology trust.*

Baseline and outcome differences are presented together in Figure 3, illustrating the relative precision of each estimate.

**Figure 3.**
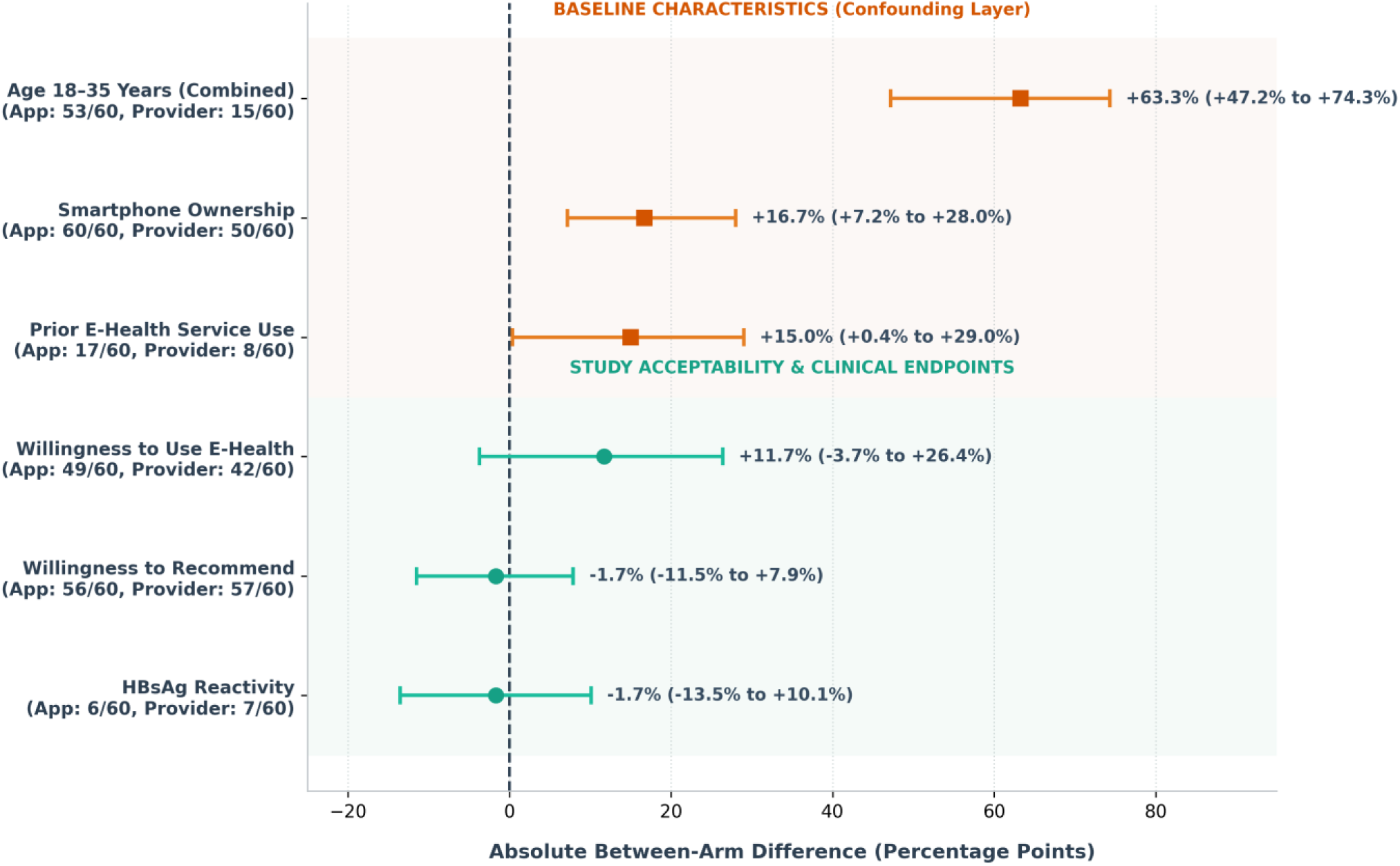
Baseline imbalances and between-arm differences in acceptance and testing outcomes. Absolute percentage-point differences between the app-guided and provider-administered arms are shown for baseline characteristics (age, smartphone ownership, prior e-health use) and for acceptance and testing outcomes (willingness, recommendation, HBsAg reactivity), with 95% confidence intervals. The figure is descriptive; confidence intervals illustrate the imprecision of the pilot estimates rather than adjusted treatment effects.

## 4. Discussion

### 4.1 Principal findings

The central tension in this pilot is that the app-guided pathway was acceptable enough to generate high willingness, yet general digital confidence did not translate into equally high ratings of the actual testing experience. At the same time, the similar HBsAg-reactive proportions across arms are too imprecise, and the design too weak, to support a claim of diagnostic equivalence. The study is therefore more informative as an implementation and usability pilot than as a diagnostic-comparison study. The E-value sensitivity analysis indicates that the observed willingness difference between arms is compatible with confounding of a magnitude plausible given the measured baseline imbalances, which reinforces rather than overturns the descriptive framing applied throughout this paper.

The observed HBsAg-reactive proportions, 10.0% (6/60) and 11.7% (7/60), are consistent with Nigeria’s high HBV burden [1–3]. As participants were a convenience sample recruited at one commercial site, these findings reflect this specific population rather than a population-representative prevalence estimate. HBsAg-reactive results in both delivery pathways underscore the importance of establishing a clear confirmatory testing and linkage-to-care process wherever community-based self-testing is implemented.

### 4.2 Self-testing evidence and the limits of the present diagnostic comparison

Self-testing programmes can expand access when instructions, result interpretation and linkage are well designed. Multi-infection self-testing work suggests that combined HBV/HCV testing is feasible in selected settings [16,17], while a much larger HIV self-testing literature shows how privacy, autonomy, kit distribution and linkage support shape uptake [18–25]. The present study adds an Ibadan implementation example, but it does not provide the evidence needed to claim diagnostic agreement. Each participant used only one pathway, there was no independent laboratory reference standard, and independent field-performance documentation for the RDT brand used was not available. A future diagnostic-comparison study should test the same participant through the intended self-testing pathway and a prespecified reference procedure, then report agreement and diagnostic accuracy with a clinically justified noninferiority or equivalence margin where appropriate [38–42].

### 4.3 Digital confidence and experienced ease of use

Three-quarters of the app-guided arm reported high baseline comfort with mobile health technology, but only 41.7% rated the specific testing tool as very or extremely easy after use. The two questions are not interchangeable measures of perceived ease of use, so a formal pre-post effect should not be inferred. Their divergence is nevertheless useful for design: familiarity with smartphones does not guarantee that users will find a health-testing workflow simple. Usability research in e-health recommends observing task completion, errors, comprehension and points of hesitation rather than relying only on anticipated acceptability [43]. Person-based and engagement frameworks likewise support iterative qualitative work in which target users shape content, sequence and interaction design [44,45]. Contemporary TAM-oriented mHealth research reinforces the need to measure experienced effort rather than assume that general technology familiarity predicts application acceptance [46,47].

The next app iteration should therefore be evaluated with task-based usability sessions before a larger trial. Useful metrics include successful independent completion of each testing step, time on task, requests for assistance, incorrect timing or interpretation of the RDT, navigation errors, comprehension of reactive/non-reactive results, and a validated usability scale. Think-aloud interviews with participants who rate the app as difficult would help identify whether the problem lies in language, procedural burden, visual hierarchy, device compatibility, connectivity, or uncertainty about interpreting the test line.

### 4.4 Privacy, cost, and trust

Privacy was selected more often than cost in this sample, which is an important local observation but not evidence that privacy is universally a larger barrier than cost for Nigerian digital health. Reviews from developing-country mHealth settings identify infrastructure, device access, affordability, skills and trust as recurring barriers [48]. Privacy and confidentiality become more consequential when an app handles stigmatised infectious-disease results or transfers health information outside the device [31,49,50]. Nigeria’s data-protection framework also creates a concrete governance context for data minimisation, lawful processing, user information and security [51].

Because the app’s data storage, transmission, account requirements, and deletion controls were not fully characterised in this study, the privacy mechanism cannot be inferred from the questionnaire response alone. The next version should make the data flow visible to users: what is collected, why it is needed, where it is stored, who can access it, how long it is retained, and how a user can delete or decline transmission (Figure 4). The low frequency of the “trust in tool accuracy” barrier should also be interpreted narrowly. It does not show that general digital trust is high; it shows only that few participants selected one accuracy-specific barrier item.

**Figure 4.**
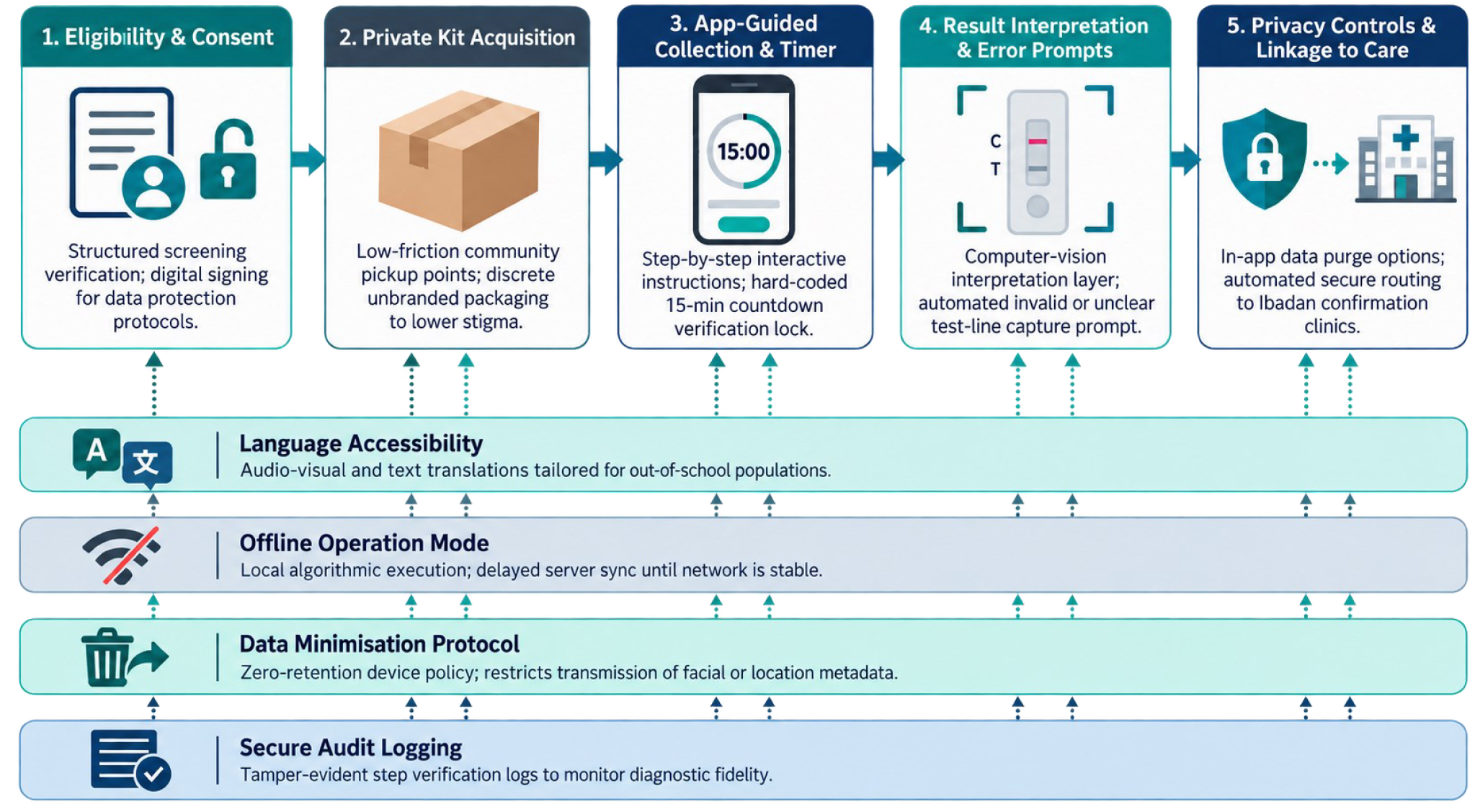
Proposed implementation pathway for privacy-aware hepatitis self-testing. A five-stage pathway from eligibility and consent through private kit acquisition, app-guided specimen collection, result interpretation, and privacy-protective linkage to care, incorporating language accessibility, offline operation, data minimisation, and secure audit logging as cross-cutting design considerations for future deployment.

Implementation studies of mobile support for HIV self-testing in southern Africa also show that users may accept digital assistance while still requiring clear, low-friction guidance through the testing process [52]. This supports treating privacy and usability as separate design domains rather than assuming that willingness alone predicts successful self-testing.

### 4.5 HCV non-detection

The screening encounter should also be interpreted within a care continuum. Global viral-hepatitis strategy emphasises diagnosis linked to prevention and treatment rather than one-off screening [53], and contemporary HBV testing recommendations similarly distinguish screening assays from subsequent clinical evaluation [54]. Future protocols should include explicit documentation of counselling, confirmation, and referral after reactive results.

No anti-HCV-reactive result was observed among 120 participants. The exact 95% CI extends to approximately 3.0%, so the result is compatible with a low but non-zero underlying prevalence. HCV epidemiology differs from HBV: risk is concentrated in specific exposure groups, and African prevalence varies substantially by country, age and population [55–57]. A small study among undergraduates in southwestern Nigeria also found low HCV antibody prevalence, illustrating that zero or near-zero detection is plausible in younger cohorts without enrichment for established HCV risk factors [58]. The present result should not be used to narrow public-health surveillance for HCV. Instead, a future study should prespecify whether HCV is a co-primary target, enrich recruitment if HCV case detection is an objective, and document the exact anti-HCV test sensitivity and specificity under the study conditions.

### 4.6 Relevance to hepatitis testing among young adults in Ibadan

Local HBV studies support the continued need for accessible testing in Ibadan and among young adult populations. HBsAg has been detected in occupational groups such as hairdressers and in student cohorts in Ibadan and elsewhere in Nigeria [59–61]. The present sample, however, was unusual in another respect: more than half of the app-guided arm held a bachelor’s degree, despite being classified as out of school, and every app-guided participant owned a smartphone. These characteristics make the sample useful for early usability testing but poorly suited to estimating uptake among digitally excluded out-of-school youths. Future recruitment should record current school status, highest completed education, employment type, income or affordability proxies, device ownership, data access and digital literacy, then sample deliberately across those strata.

### 4.7 Methodological implications for a definitive study

A definitive evaluation should separate three questions that were combined in this pilot. First, diagnostic performance requires a paired or reference-standard design. Second, service-delivery effectiveness requires randomised or otherwise well-controlled allocation, with prespecified outcomes such as completed testing, correct test execution, result comprehension and linkage after a reactive result. Third, technology acceptance requires validated, prospectively specified measures rather than retrospective mapping of questionnaire items to TAM and UTAUT. These components can be combined in one protocol, but their estimands are different.

For acceptance outcomes, random allocation would reduce the age and digital-readiness imbalance seen here. If randomisation is not feasible, participant-level multivariable modelling or prespecified stratification should address age, sex, education, smartphone ownership, prior e-health use and relevant hepatitis risk factors. For feasibility outcomes, progression criteria should be defined before recruitment, for example the proportion completing the app workflow without assistance, the proportion obtaining an interpretable RDT, the time required, data-completeness thresholds and the proportion of reactive participants successfully linked to confirmatory care. Sample size for the definitive trial should then be based on the primary estimand and a clinically meaningful effect or equivalence margin rather than on retrospective power.

### 4.8 Strengths and limitations

This pilot has several strengths: both arms used the same rapid-test brand, holding assay type constant while the delivery pathway varied; the study was conducted in a real-world informal commercial setting; and it captured both pre-use expectations and post-use experience in the app-guided arm. Key limitations include convenience sampling at a single site with non-random allocation, which produced a marked baseline imbalance in age, smartphone ownership, and prior e-health use that limits causal interpretation of between-arm acceptance comparisons; TAM/UTAUT constructs measured with single, non-validated items rather than multi-item scales; and the absence of independent performance-validation data for the rapid test kits used. These limitations are addressed through the descriptive framing, sensitivity analysis, and forward-looking recommendations presented throughout this paper.

## 5. Conclusion

This pilot shows that app-guided hepatitis rapid testing can be implemented among digitally connected out-of-school young adults in a commercial setting in Ibadan, and that stated willingness to use the approach is high. It does not establish diagnostic equivalence with provider-administered testing. The wide confidence interval around the HBsAg difference, the lack of a paired reference standard and the substantial baseline imbalance require a narrower interpretation.

The most useful design signal is experiential. High general comfort with mobile health did not translate into equally high ease ratings after participants used the hepatitis-testing workflow, and privacy was selected by roughly one-third of participants. Before scaled deployment, the application would benefit significantly from task-based user testing, privacy-by-design review and explicit documentation of confirmatory testing and within-app linkage to care pathway. A larger study should randomise the delivery pathway or use an appropriate paired diagnostic design, prespecify its primary estimand and precision target, and use validated acceptance and usability measures.

## Abbreviations

AI: artificial intelligence
CI: confidence interval
HBsAg: hepatitis B surface antigen
HBV: hepatitis B virus
HCV: hepatitis C virus
LMIC: low- and middle-income country
mHealth: mobile health
RDT: rapid diagnostic test
TAM: Technology Acceptance Model
UTAUT: Unified Theory of Acceptance and Use of Technology

## Acknowledgements

The authors thank the participants and field personnel involved in recruitment and testing at the Iwo Road study site. The authors also acknowledge the Iwo Road market association leaders and communities for their support for digital-health research. Also acknowledged is the Skrind Biotech technical team for their continued support and scientific capacity building.

## Author Contributions (CRediT)

A.O.A: Conceptualisation, Methodology, Formal Analysis, Writing - Review & Editing, Supervision. R.M.Y: Investigation, Field Activity, Methodology, Data Curation, Writing - Original Draft. A.R.O: Validation, Field Activity Coordination, Writing - Review & Editing. N.N.A: Methodology, Formal Analysis, Writing - Review & Editing. A.A: Validation, Writing - Review & Editing, Supervision. All authors read the final manuscript and approved before submission.

## Ethical Approval and Informed Consent

The study received ethical clearance from the Oyo State Health Research Ethics Committee (HREC), Ibadan, Nigeria with approval number: NHREC/OYO/SHRIEC/10/11/22. Written informed consent was obtained from all participants before enrolment and testing, and identifying information was replaced with unique participant codes after data collection.

## AI Use Statement

Generative AI tools were used to assist with language refinement, literature organisation, and document formatting during manuscript preparation. All scientific content, citations, and statistical statements were verified by the authors against the underlying data and primary sources. The authors take full responsibility for the final content of this manuscript.

## Data Availability

The de-identified participant-level dataset underlying this study are not publicly available and will not be shared. Participants were recruited from a community-based, informal-sector population in relation to a stigmatised health condition, and the original informed consent and institutional ethical approval did not include provision for third-party data sharing or open-data deposition. All aggregated results necessary to interpret and evaluate the findings reported in this manuscript are presented within the article and its tables. Requests for methodological clarification may be directed to the corresponding author.

